# Surgical practices followed during containment, management and resolution of gastrointestinal fistulas. Results of a multicontinent, multinational, multicentric study

**DOI:** 10.1101/2022.06.18.22276589

**Authors:** Humberto Arenas Márquez, María Isabel Turcios Correia, Juan Francisco García, Roberto Anaya Prado, Arturo Vergara, Jorge Luis Garnica, Alejandra Cacho, Daniel Guerra, Miguel Mendoza Navarrete, Sergio Santana Porbén

## Abstract

**Introduction:** The “Fistula Day” multicontinent, multinational, multicentered project has revealed a 14.7 % mortality rate in patients assisted for gastrointrestinal fistulas (GIF) in Latin American and European hospitals. Mortality associated with GIF might be explained for the clinical-surgical condition of the patient, the operational characteristics of the hospital, and the surgical practices locally adopted in the contention, treatment and resolution of GIF.

**Objective:** To assess the influence of surgical practices adopted in the hospital upon GIF outcomes.

**Study design:** Cohort-type study. Three cross-sectional examinations were done during the completion of the exercises of the “Fistula Day” project: on admission in the study serie, and at 30 and 60 days after admission.

**Study serie:** One hundred seventy seven patients (*Males*: 58.2 %; *Average age*: 51.0 ± 16.7 years; *Ages* _≥_ *60 years*: 36.2 %) assisted in 76 hospitals of Latin America (13 countries) and Europe (4).

**Methods:** Surgical practices adopted in the management of GIF were documented such as the use of computerized axial tomography (CAT) and oral ingestion of contrast for examination of the fistula path, the use of open abdomen and devices for temporary closure of the abdominal wall, the administration of somatostatin and analogs for promoting the closure of the fistula, reoperation for fistula closure, and admission in the ICU.

**Results:** Usage rate of surgical practices was as follows: *CAT + oral use of contrast*: 39.5 %; *Use of open abdomen*: 31.1 %; *Use of somatostatin and analogs*: 22.6 %; *Admission in the hospital ICU*: 31.6 %; and *Surgery for GIF closure*: 33.9 %; respectively. Surgical practices were more frequently used in the treatment and containment of enteroathmosferic fistulas (EAF). Surgical practices adopted by participating hospitals did not imply a higher rate of GIF closure, but were associated instead with a higher mortality and prolongation of hospital stay. Conduction of surgical practices was independent from the guidelines followed by the medical teams in the management of GIF. Availability of surgical practices, and access of medical teams to them, were independent from the operational characteristics of the surveyed hospital. It is to be noticed the existence of a hospital unit dedicated to intestinal failure translated to a lower use of the techniques for open abdomen and temporary closure of the abdominal wall, which, in turn, translated to a higher likelihood of GIF spontaneous closure.

**Conclusions:** Currently, the adoption of surgical practices for containment and resolution of GIF does not result in a higher GIF closure rate. It is likely the existence of a hospital unit specialized in the management of intestinal failure might bring about a higher rate of non-surgical closure of GIF.

## INTRODUCTION

The “Fistula Day” Project is an initiative aimed to the improvement of containment and treatment of gastrointestinal fistulas (GIF) in Latin America (LATAM) hospitals. To such end, the project foresees the regular conduction of surveys with the purpose of revealing those surgical practices related with the best outcomes in the treatment of GIF.

GIF usually affect between 4 – 20 % of the operated patients.^1-2^ However, these estimates might vary according with the characteristics of the health institution, and the volume and complexity of the surgeries completed in a working year. Containment, treatment and resolution (non-surgical | surgical) of GIF imply prolonged hospital stays, addititional quotas of surgical-medical actions, and increased health costs. However, and in spite of the therapeutic efforts, mortality associated with GIF might be as high as 80 % of the affected patients.^3-4^

Previous publications have explored the impact of the clinical-surgical condition of the patient and the operational characteristics of the hospital containing and care for him | her, upon the indicators of the evolution and ultimate outcome of GIF, namely, survival, prolongation of hospital stay, and spontaneous closure (also read as non-surgical | conservative) of the fistula, respectively.^5-6^ In the first of these works presence of an entero-athmospheric fistula (EAF) was associated with a higher mortality, while hospital stay was dependent upon the type of surgery initially performed, location of fistula, and nutritional status of the patient (as estimated from calf circumference at admission in the study serie).^5^ However, none of the characteristics of the patient determined the spontaneous closure of the fistula.^5^

In a follow-up article, the number of hospital beds emerged as the best predictor of the survival of the GIF patient, prolongation of hospital stay, and a higher rate of spontaneous closure of the fistula;^6^ but other operational characteristics of the hospital, such as the existence of a unit specialized in intestinal failure and the number of patients assisted during one month, might also influence (albeit marginally) upon evolution of GIF.

Findings of the “Fistula Day” documented so far might point towards the nature and content of surgical practices conducted upon the patient during containment and treatament of GIF. Regarding the aforementioned, treatment and resolution of GIF have called the attention of groups of experts and profesional societies, and the “ASPEN-FELANPE Clinical Guidelines: Nutrition support of adult patients with enterocutaneous fistula” might be mentioned among the guidelines drafted for this purpose.^7-8^ Identification of the source of the fistula, containment of the surgical damage, and externalization of the fistula stoma might be actions initially recommended. Further actions in the treatment of GIF might include the best assessment on their likely spontaneous closure, and the place of medications such as glues and somatostatin (Octeotride®©, Sandoz, Switzerland).

Given all the aforementioned, completion of the “Fistula Day” was a unique opportunity to record the surgical practices locally conducted in the participating hospitals to achieve the closure of the fistula, first; and to assess their impact upon the condition of the patient upon discharge and prolongation of hospital stay.

## MATERIAL AND METHOD

The design of the “Fistula Day” Project has been previously described.^5-6^ Briefly, hospitals eventually included in the project were invited to submit the demographical, clinical, sanitary, surgical and nutritional data of patients complicated with GIF between the months of May of 2019 and July of 2019 (both included) in three consecutive surveys.^5^ Hospitals also submitted data on their operational characteristics, the number of hospital beds and the number of patients assisted | treated for GIF in a month-work among them.^6^ Participating hospitals were also surveyed on the presence within the institution’s organigram of an Intensive Care Unit (ICU), a multidisciplinary unit dedicated to Clinical Nutrition (MDUCN), and a unit dedicated to the treatment of intestinal failure and/or postoperatory fistula (IFU). In addition, hospitals were asked about the expertise of the acting physician on the treatment of intestinal leakages | fistulas.^6^

The design of the “Fistula Day” Project foresaw the recording of surgical practices adopted in the containment, treatment and eventual resolution of GIF, the use of the open abdomen technique, administration of somatostatin (or analogs of the hormone in its defect) for the non-surgical closure of the fistula, and admission to a hospital ICU among them.

### Data processing and statistical-mathematical analysis of the results

Data submitted by the hospitals involved in the project were entered into an on line application built upon *RedCap*®©^*^ (University of Vanderbilt, United States). The R program for statistical management and analysis (R Core Team 2018 version 3.5.0, United States) was used for debugging, preparing and processing data collected during the surveys of the “Fistula Day”. Data were reduced down to absolute | relative frequencies and percentages according with the type of the variable and the objective of the statistical analysis.

Condition of the patient (Alive/Deceased) upon discharge, hospital stay (Yes/No) and spontaneous closure of GIF (Yes/No) were assumed as the project outcomes at 30 and 60 days of the hospìtal’s admission in the “Fistula Day”. Nature and strength of the associations between the outcomes of the “Fistula Day” on one hand, and the identified surgical practices, on the other; were examined by means of appropriate statistical *tests* in accordance with the type of the variable. Differences arising between cohorts of patients regarding the selected predictor were assessed by means of the log-rank test based in the chi-squared distribution.^7^ A level lower than 5 % was used in all the instances to denote the finding as significant.

### Treatment of missing data

Data missing during follow-up of the patient were replaced with the observation recorded in the previous cross-sectional examination according with the LOCF (“Last Observation Carried Forward”) method.

### Intention-to-treat

Data gathered during the “Fistula Day” were analyzed according with the “Intention to treat” principle in order to keep fixed the size of the cohort.^8^

### Ethical considerations

The protocol followed by local surveyors during the “Fistula Day” was drafted in accordance with the “Good Clinical Practices” guidelines.^9^ Identity and rights of the surveyed patients were protected at all times.^10^ Patients (and by extension their caregivers) were informed about the purposes of the research, and the non-invasive nature of the procedures. Collected data were adequately preserved in order to ensure anonymity and confidentiality. Aggregated data were used solely for interpretation of the results and realization of statistical inferences. Informed consent was obtained from the GIF patient before inclusion in the cohort. Local conduction of the activities foreseen in the “Fistula Day” was authorized and supervised by the hospital Committees of Ethics after presentation, review and approval of the research protocols.

The researchers entrusted with the conduction of the “Fistula Day” Project presented the protocol “Current status of the postoperative fistula of digestive tract; multicentric, multinational study. DAY OF THE FISTULA” before the Ethics Committee of the San Javier Hospital (city of Guadalajara, State of Jalisco, México) for review and approval. A ruling was emitted on April 11^th^, 2018 by Dr. Eduardo Razón Gutiérrez, acting Director of the Ethics Committee, with the approval of the research protocol and the authorization for the conduction of the “Fistula Day” Project.

## RESULTS

### Main characteristics of the participating hospitals

Seventy-six hospitals from 17 countries participated in the “Fistula Day” activities.^6^ Thirteen of the participating countries were from LATAM. Sixty of the hospitals were mexicans. Specialties hospitals prevailed (at least numerically). Most of the hospitals assisted between 1 – 2 GIF patients in a month-work. Participating hospitals distributed evenly regarding the number of beds. Most of the hospitals counted with a unit specialized in the delivery of intensive care (ICU). In addition, three-quarters of the hospitals had a multidisciplinary unit dedicated to Clinical and hospital nutrition. On the contrary, a unit dedicated to the treatment of intestinal failure and/or postoperatory fistulas was only present in one-quarter of the hospitals. Expertise in GIF management of the acting physician was rated between “Expert” and “High” in one-third part of the hospitals.

### Main characteristics of the surveyed patients

One-hundred seventy-seven patients were surveyed during the “Fistula Day” exercises.^5^ Men prevailed over women accounting for 58.2 % of the size of the study serie. Average age was 51.0 ± 16.7 years. Subjects with ages _≥_ 60 years represented 36.2 % of the studied cases. Fifty nine-point-six percent of the patients accumulated between 0 – 30 days of hospital stay at admission in the study serie. A diagnosis of cancer had been made in 27.7 % of the patients. Enterocutaneous fistula (ECF) was the prevailing type of fistula in the study serie. Almost 60 % of the GIF showed an output < 500 mL.day^-1^. Small bowel and colon were the dominant locations as origins of GIF. Half-plus-one of the GIF was diagnosed after the first 5 days of the primary surgery. In addition, 60.5 % of GIF originated after an emergency surgery.

### Main results of the “Fistula Day” Project

On conclusion of the study the indicators of the evolution of GIF behaved as follows: *Mortality*: 14.7 %; *Prolonged hospitalization*: 46.3 %; and *Spontaneous closure of fistula*: 36.2 %.

### Surgical practices adopted in patients with gastrointestinal fistulas

Diagnosis of GIF by means of CAT + oral ingestion of contrast was made only in 39.5 % of the patients. The type of fistula did not influence upon the use of CAT + oral ingestion of contrast: EAF: 43.5 % *vs*. ECF: 37.4 % (Δ = +6.1 %; p > 0.05).

Open abdomen technique had been used as primary method for containing GIF in 31.1 % of the patients. Temporary closure of the wall was more used in the treatment of EAF: EAF: 54.8 % *vs*. ECF: 18.3 % (Δ = +36.5 %; p < 0.05). Bogotá bag (58.2 % of the patients with temporary closure of the abdominal wall), and the VAC system for vacumm aspiration (41.8 %) were the most used devices for closing the abdominal wall temporarily, regardless the type of GIF.

A therapeutic trial with somatostatin (*Octreotide*®©, Sandoz, Switzerland) for closure of GIF was made in 22.6 % of the patients. The hormone was administered preferibly subcutaneously (67.5 % of the instances) between 3 – 10 days (70.0 %). Treatment with somatostatin concentrated in EAF patients: EAF: 41.9 % *vs*. ECF: 12.2 % (Δ = +29.7 %; p < 0.05).

Thirty-one-point-six percent of GIF patients required admission in a hospital ICU. Seventy-three-point-two percent of them accumulated up to 20 days of stay in the ICU. EAF consumed a higher proportion of ICU admissions: EAF: 56.5 % *vs*. ECF: 18.3 % (Δ = +38.2 %; p < 0.05). Half-plus-one of the GIF patients required mechanical ventilation during ICU admission. Again, EAF patients required a higher quota of mechanical ventilation: EAF: 37.1 % *vs*. ECF: 8.7 % (Δ = +29.4 %; p< 0.05).

On admission of the patient in the study, a surgery for closure of GIF had been completed in 33.9 % of them. Rate of surgical closure of GIF was (at least numerically) higher in EAF patients: EAF: 45.2 % *vs*. ECF: 27.8 % (Δ = +17.4 %; p > 0.05).

### Impact of surgical practices upon survival of the patient

Of the examined surgical practices, only ICU admission influenced upon survival of GIF patients, but the effect was paradoxical: survival was higher among patients not admitted to an ICU: Alive: *ICU admission*: 76.8 % vs. *No ICU admission*: 89.3 % (Δ = -12.5 %; χ^2^ = 4.75; p < 0.05). The remaining surgical practices did not determine a higher survival of the GIF patient: Alive: *CAT use + oral constrast*: 85.7 % vs. *No CAT use*: 85.0 % (Δ = +0.7 %; p > 0.05); *Use of open abdomen*: 81.8 % vs. *No use of open abdomen*: 86.9 % (Δ = -5.1 %; p > 0.05); *Treated with somatostatin*: 82.5 % vs. *No treated*: 86.1 % (Δ = -3.6 %; p > 0.05); and *Reoperation for GIF resolution*: 81.7 % vs. *No reoperation*: 87.2 % (Δ = -5.5 %; p > 0.05).

The impact of surgical practices upon survival of the GIF patient was assessed in parallel with the type of fistula. In all the instances, type of fistula determined survival of the patient instead of the adopted surgical practice. Hence, the odds for survival in ECF patients were always higher: *Use of CAT + oral ingestion of contrast*: OR_Type fistula_ = 2.055 [IC 95 %: 1.164 – 3.629; p < 0.05] *vs*. OR_Treatment_: 1.505 [IC 95 %: 0.787 – 2.878; p > 0.05]; *Use of open abdomen*: OR_Type fistula_ = 2.289 [IC 95 %: 1.379 – 3.797; p < 0.05] *vs*. OR_Treatment_: 1.339 [IC 95 %: 0.745 – 2.402; p > 0.05]; *Treatment with somatotastin*: OR_Type fistula_ = 2.211 [IC 95 %: 1.310 – 3.733; p < 0.05] *vs*. OR_Treatment_: 1.442 [IC 95 %: 0.747 – 2.783; p = 0.0575]; *ICU admission*: OR_Type fistula_ = 2.722 [IC 95 %: 1.639 – 4.521; p < 0.05] *vs*. OR_Treatment_: 1.067 [IC 95 %: 0.615 – 1.815; p > 0.05]; and *Reoperation for resolution of GIF*: OR_Type fistula_ = 1.666 [IC 95 %: 1.073 – 2.589; p < 0.05] *vs*. OR_Treatment_: 1.398 [IC 95 %: 0.819 – 2.386; p < 0.05]; respectively.

Figure 1 shows the impact of surgical practices upon survival of the GIF patient when cases were analyzed as a cohort. Of the surgical practices considered, only admission in the ICU implied a lower number of survivors in each time point of the cohort of cases: *Second time point*: No ICU Admission: 91.7 % vs. ICU Admission: 83.9 % (Δ = +7.8 %); *Third time point*: No ICU Admission: 89.3 % vs. ICU Admission: 76.8 % (Δ = +12.5 %; χ^2^ = 4.305; p < 0.05; log-rank test). Given the paucity of data, no attempt was made to assess the influence of the type of GIF fistula upon the mortality of the cohort of cases.

**Figure 1.**
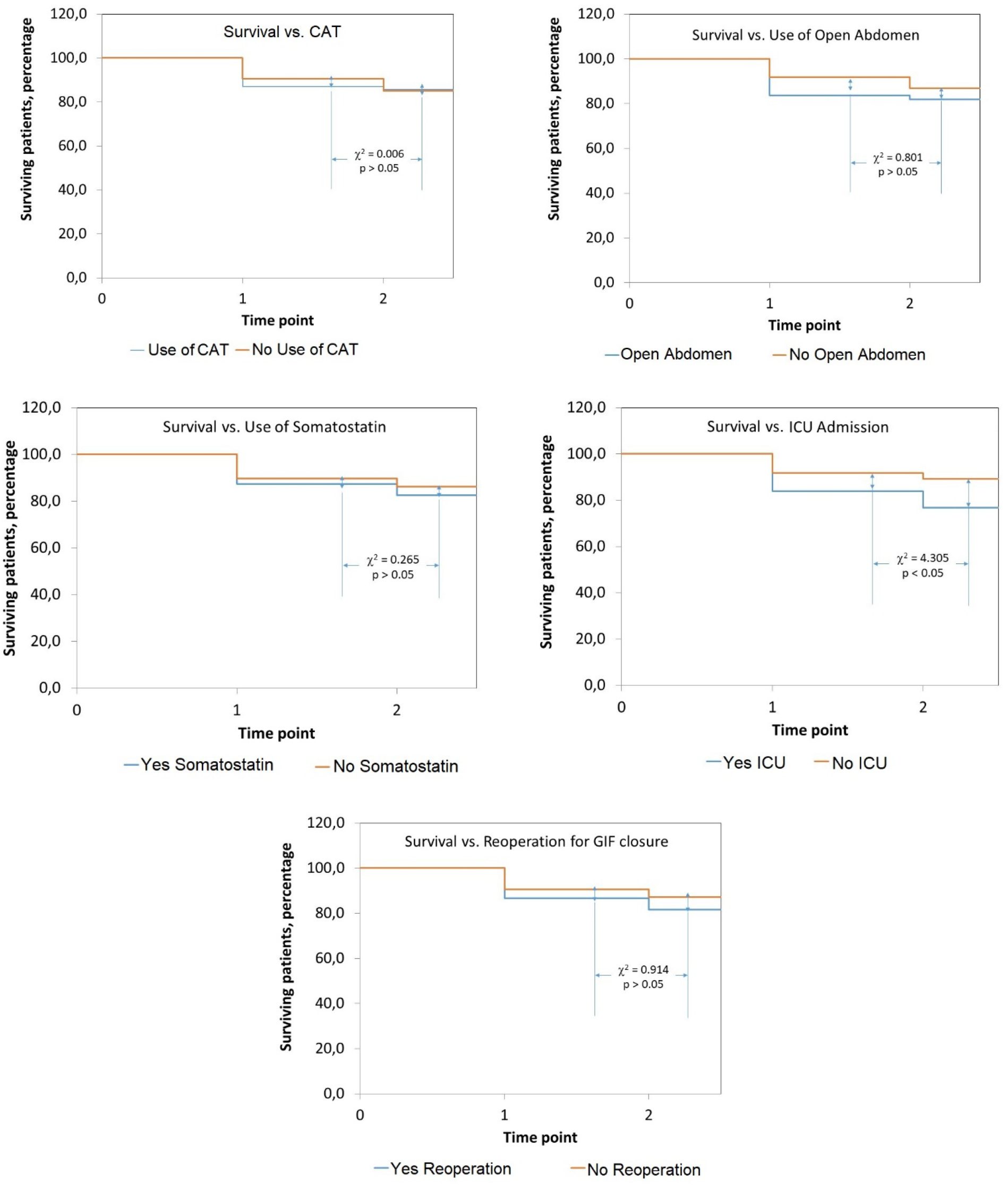
Impact of assessed surgical practices upon survival of patients with gastrointestinal fistulas. The study serie was dessagregated into the corresponding cohorts. Source: Records of the study. Size of the study serie: 177.

### Impact of surgical practices upon hospitalization of the patient

The impact of the surgical practices upon timely hospital discharge (that is: without requiring prolongation of the hospital stay) was also examined. However, use of the open abdomen (Yes: 40.7 % *vs*. No: 57.7 %; Δ = -17.0 %; χ^2^ = 4.340; p < 0.05), admission in the ICU (Yes: 37.5 % *vs*. No: 59.5 %; Δ = -22.0 %; χ^2^ = 7.433; p < 0.05), and reoperation for closure of the fistula (Yes: 28.3 % *vs*. No: 71.0 %; Δ = -42.7 %; χ^2^ = 23.328; p < 0.05) translated to the prolongation of the hospital stay.

Impact of surgical practice upon timely hospital discharge was assessed in parallel with the type of fistula. Type of fistula did not determine a timely hospital stay, while 4 of the surgical practices had a neutral effect: *Use of CAT + Oral ingestion of constrast*: OR_Type fistula_ = 1.338 [IC 95 %: 0.890 – 2.012; p > 0.05] *vs*. OR_Treatment_: 0.748 [IC 95 %: 0.466 – 1.200; p > 0.05]; *Use of open abdomen*: OR_Type fistula_ = 1.000 [IC 95 %: 0.689 – 1.449; p > 0.05] *vs*. OR_Treatment_: 0.959 [IC 95 %: 0.613 – 1.501; p > 0.05]; *Use of somatostatin*: OR_Type fistula_ = 0.957 [IC 95 %: 0.655 – 1.397; p > 0.05] *vs*. OR_Treatment_: 1.000 [IC 95 %: 0.615 – 1.627; p > 0.05]; and *Admission in the ICU*: OR_Type fistula_ = 1.000 [IC 95 %: 0.689 – 1.450; p > 0.05] *vs*. OR_Treatment_: 0.809 [IC 95 %: 0.517 – 1.265; p > 0.05].

Coming to this point, it is to be noticed the effect of reoperation upon the closure of fistula. Thus, timely hospital discharge was very likely in ECF patients explained mainly because of the presence of this type of fistula: OR_Type fistula_ = 1.69 [IC 95 %: 1.12 – 2.56; p < 0.05]; while reoperation implied a disminished likelihood of timely hospital discharge: OR_Treatment_: 0.521 [IC 95 %: 0.318 – 0.852; p < 0.05].

Figure 2 shows the impact of surgical practices upon prolongation of hospital stay of the cohorts of cases. Of the surgical practices considered, only reoperation for surgical closure of GIF implied prolongation of hospital stay in each time point of the cohort of cases: *Second time point*: No Reoperation: 27.4 % vs. Reoperation: 51.7 % (Δ = -24.3 %); *Third time point*: No Reoperation: 7.7 % vs. Reoperation: 20.0 % (Δ = -12.3 %; χ^2^ = 6.470; p < 0.05; log-rank test). Given the paucity of data, no attempt was made to assess the influence of the type of fistula upon prolongation of hospital stay in the cohort of cases.

**Figure 2.**
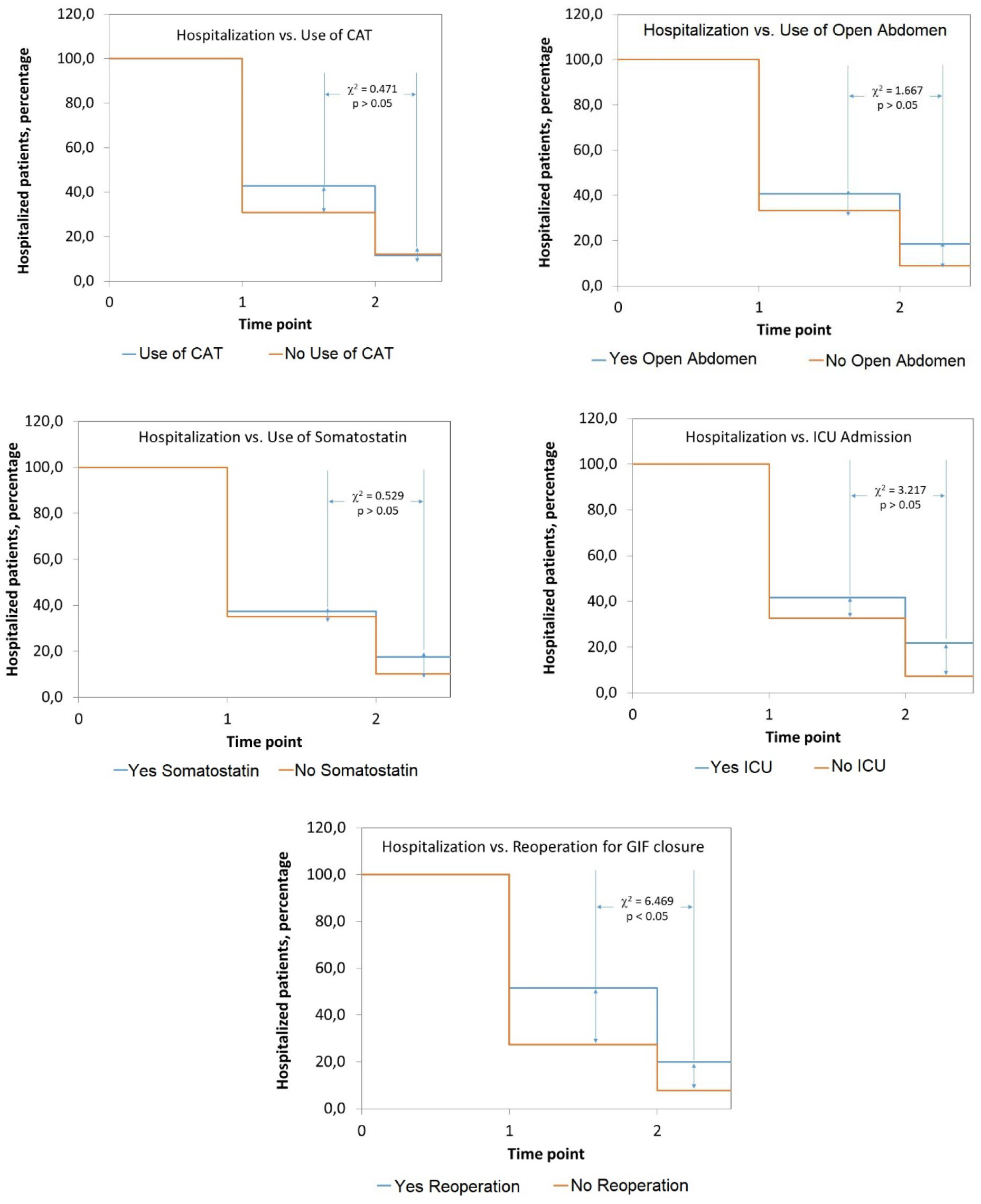
Impact of assessed surgical practices upon hospital stay of patients with gastrointestinal fistulas. The study serie was dessagregated into the corresponding cohorts. Source: Records of the study. Size of the study serie: 177.

In this point of the discussion, it is to be noticed admission in the UCI translated (albeit marginally) to a prolonged hospital stay: Hospitalized patients: *Second time point*: No admission in the ICU: 32.8 % vs. Admission in the ICU: 41.8 % (Δ = -9.0 %); *Third time point*: No admission in the ICU: 7.4 % vs. Admission in the ICU: 21.8 % (Δ = -14.4 %; χ^2^ = 3.217; p = 0.073; test de log-rank): an interesting finding given the heterogeneity of the cohort. Similar to the aforementioned in the preceeding sections, data paucity impeded assessing the influence of the type of the fistula upon prolongation of the hospital stay in the cohort of cases.

### Impact of the surgical practices upon the succesful surgical closure of the gastrointestinal fistula

Succesful closure (that is: without further refistulization) of the fistula is the ultimate goal of the surgical practices adopted in the patient. Twenty-five refistulization events were recorded during the window of observation of the study. Use of open abdomen (Yes: 22.2 % vs. No: 11.5 %; Δ = +10.7 %; χ^2^ = 4.201; p < 0.05) and reoperation for surgical closure of the fistula (Yes: 23.3 % vs. No: 9.1 %; Δ = +13.9 %; χ^2^ = 6.346; p < 0.05) were associated with refistulization: patients subjected to these practices showed a higher frequency of refistulizations. Remaining practices had a neutral effect.

Succesful closure of fistula was assessed in parallel with the surgical practices described in the preceeding sections. Administration of CAT + oral use of constrast and admission in the ICU translated to higher likelihood of succesful closure, even in spite of the type of fistula: *CAT + Oral use of constrast*: OR_Type fistula_ = 1.826 [IC 95 %: 1.000 – 3.333; p < 0.05] *vs*. OR_Treatment_: 1.774 [IC 95 %: 0.894 – 3.520; p < 0.05]; *Admission in the ICU*: OR_Type fistula_ = 1.947 [IC 95 %: 1.146 – 3.306; p < 0.05] *vs*. OR_Treatment_: 1.698 [IC 95 %: 0.909 – 3.169; p < 0.05].

Remaining practices only had a neutral effect on the possibility of a succesful closure of the fistula, while the effect of the type of fistula prevailed in every moment: the possibility of a succesful closure was higher in ECF: *Use of open abdomen*: OR_Type fistula_ = 1.826 [IC 95 %: 1.000 – 3.333; p < 0.05] *vs*. OR_Treatment_: 1.774 [IC 95 %: 0.894 – 3.521; p > 0.05]; *Use of somatostatin*: OR_Type fistula_ = 1.981 [IC 95 %: 1.140 – 3.442; p < 0.05] *vs*. OR_Treatment_: 1.721 [IC 95 %: 0.857 – 3.457; p < 0.05]; and *Reoperation for surgical closure of fistula*: OR_Type fistula_ = 2.821 [IC 95 %: 1.579 – 5.040; p < 0.05] *vs*. OR_Treatment_: 1.052 [IC 95 %: 0.570 – 1.941; p > 0.05]; respectively.

Figure 3 shows the impact of the surgical practices upon refistulization of the cohort of cases. Use of open abdomen and reoperation for surgical closure of GIF were the practices associated with refistulization of the patient: Non-refistulized patients: Use of open abdomen: *Second time point*: No Open Abdomen: 95.9 % vs. Yes Open Abdomen: 85.2 % (Δ = +10.7 %); *Third time point*: No Open Abdomen: 89.4 % vs. Yes Open Abdomen: 77.8 % (Δ = +11.6 %; χ^2^ = 4.062; p < 0.05; log-rank test); Reoperation for surgical closure of GIF: *Second time point*: No Reoperation: 94.9 % vs. Reoperation: 88.3 % (Δ = +6.6 %); *Third time point*: No Reoperation: 90.6 % vs. Reoperation: 76.7 % (Δ = +13.9 %; χ^2^ = 5.860; p < 0.05; log-rank test). Given the paucity of data, no attempt was made to assess the influence of the type of fistula upon prolongation of hospital stay of the cohort of cases.

**Figure 3.**
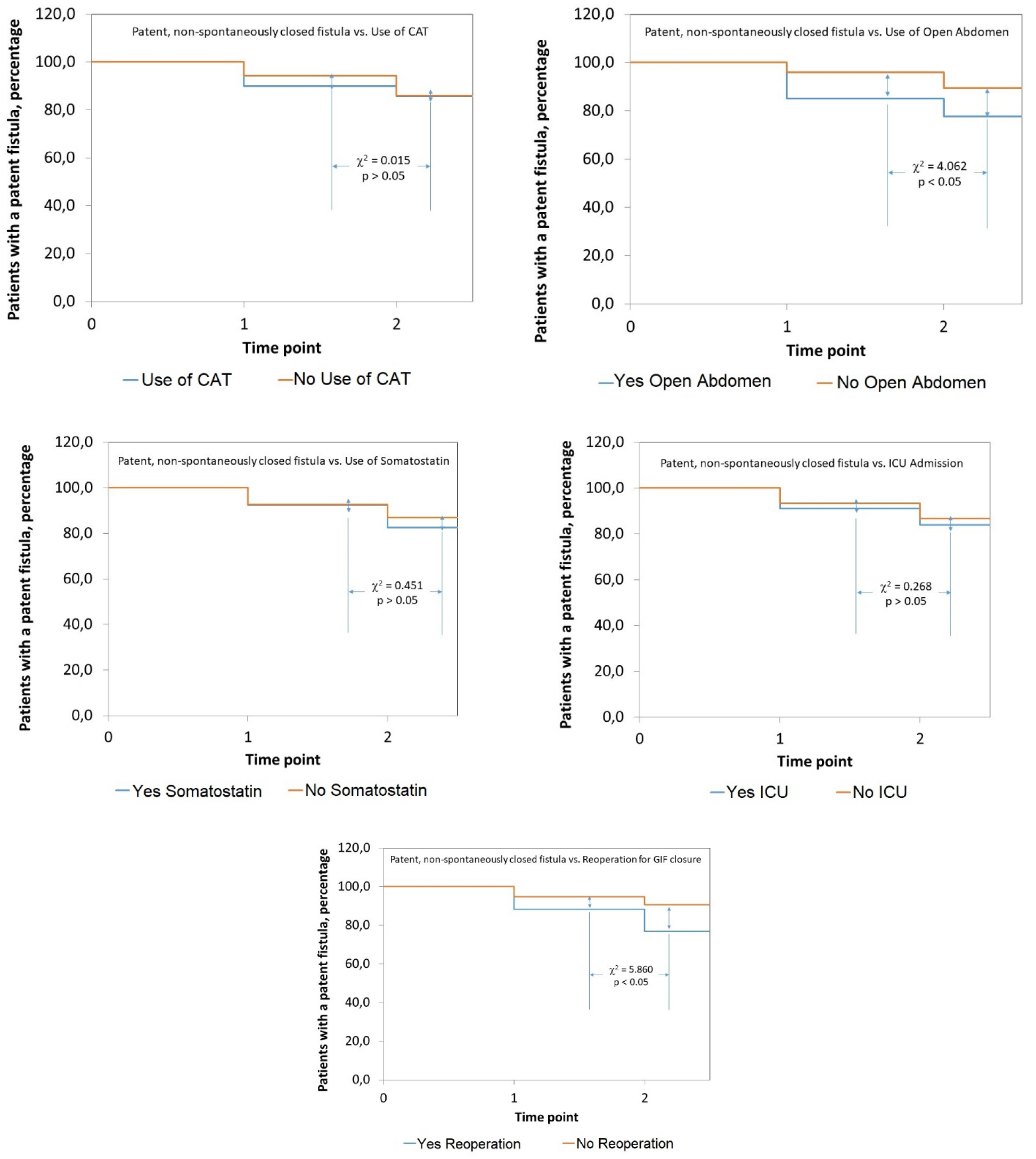
Impact of assessed surgical practices upon refistulization of patients with gastrointestinal fistulas. The study serie was dessagregated into the corresponding cohorts. Source: Records of the study. Size of the study serie: 177.

### Influence of the operational characteristics of the hospital upon the surgical practices followed in the resolution of gastrointestinal fistulas

Table 2 shows the influence of the operational characteristics of the participating hospital upon the surgical practices followed in the containment and resolution of GIF. Position of the hospital within the health system did not influenced upon the conduction of the surveyed surgical practices.

**Table 1.** Surgical practices conducted in the patients assisted for gastrointestinal fistulas in the hospitals participating in the activities of the “Fistula Day” Project. Unless otherwise indicated, results correspond with percentages of the size of the study serie. Legend: CAT: Computed Axial Tomography. TPN: Total Parenteral Nutrition. ICU: Intensive Care Unit.

| Surgical practice | Enteroatmospheric fistulas | Enterocutaneous fistulas | All the fistulas |
| --- | --- | --- | --- |
| Size | 62 | 115 | 177 |
| • ¿Was CAT + oral ingestion of contrast used? | Yes; 43.5<br>No: 56.5 | Yes; 37.4<br>No: 62.6 | Yes: 39.5<br>No: 60.5 |
| • ¿Was temporary closure of abdominal wall used for containing the fistula? | Yes: 54.8<br>No: 32.3<br>Non declared: 12.9 | Yes: 18.3<br>No: 80.9<br>Non declared: 0.9 | Yes: 31.1<br>No: 63.8<br>Non declared: 5.1 |
| • Device used <sup>¶</sup> | Bogotá bag: 67.6<br>VAC System: 47.1<br>Mesh: 11.8<br>Wittman patch: 2.9<br>Non declared: 2.9 | Bogotá bag: 42.9<br>VAC System: 33.3<br>Mesh: 14.3<br>Wittman patch: 4.8<br>Non declared: 57.2 | Bogotá bag: 58.2<br>VAC System: 41.8<br>Mesh: 12.7<br>Wittman patch: 3.6<br>Non declared: 23.6 |
| • ¿Was a therapeutic trial done with somatostatin (Octeotride)? | Yes: 41.9<br>No: 59.1 | Yes: 12.2<br>No: 87.8 | Yes: 22.6<br>No: 77.4 |
| • ¿Was route was used? <sup>§</sup> | Subcutaneous: 61.5<br>IV: 34.6<br>As part of TPN: 3.8 | Subcutaneous: 78.6<br>IV: 21.4<br>As part of TPN: 0.0 | Subcutaneous: 67.5<br>IV: 30.0<br>As part of TPN: 2.5 |
| • ¿How many days of treatment? <sup>§</sup> | 3 – 10 days: 65.4<br>11 – 20 days: 34.6 | 3 – 10 days: 78.6<br>11 – 20 days: 21.4 | 3 – 10 days: 70.0<br>11 – 20 days: 30.0 |
| • Admission to the ICU | Yes: 56.5<br>No: 43.5 | Yes: 18.5<br>No: 81.7 | Yes: 31.6<br>No: 68.4 |
| • Length of stay in the ICU | 1 – 3 days: 8.6<br>4 – 10 days: 34.3<br>11 – 20 days: 25.7<br>21 – 30 days: 0.0<br>> 30 days: 4.0<br>Non declared: 28.6 | 1 – 3 days: 23.8<br>4 – 10 days: 38.1<br>11 – 20 days: 19.0<br>21 – 30 days: 0.0<br>> 30 days: 0.0<br>Non declared: 19.0 | 1 – 3 days: 14.3<br>4 – 10 days: 35.7<br>11 – 20 days: 23.2<br>21 – 30 days: 0.0<br>> 30 days: 1.8<br>Non declared: 25.0 |
| • Mechanical ventilation | Yes: 37.1<br>No: 63.1 | Yes: 8.7<br>No: 91.3 | Yes: 58.9<br>No: 41.1 |
| • Reoperation for closure of the fistula | Yes: 45.2<br>No: 54.8 | Yes: 27.8<br>No: 72.2 | Yes: 33.9<br>No: 66.1 |

<sup>¶</sup> Presented percentages correspond with patients in whom a device for closure of the abdominal wall was used.
<sup>¶</sup> Several devices were used at the same time in several patients.
<sup>§</sup> Presented percentages correspond with patients in whom somatostatin (or its analogs) was used.
Source: Records of the study.
Size of the study serie: 177.

**Table 2.**
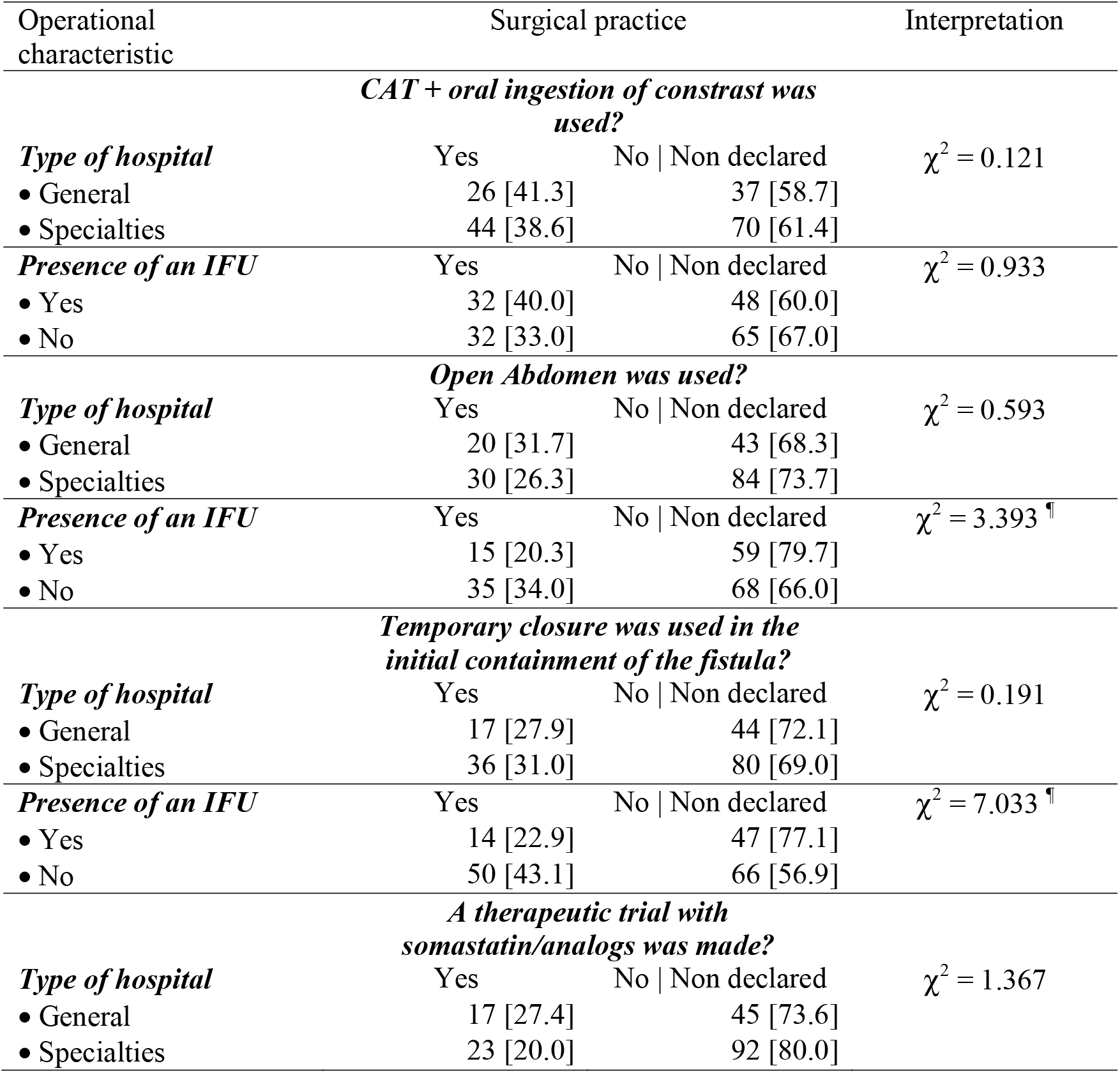
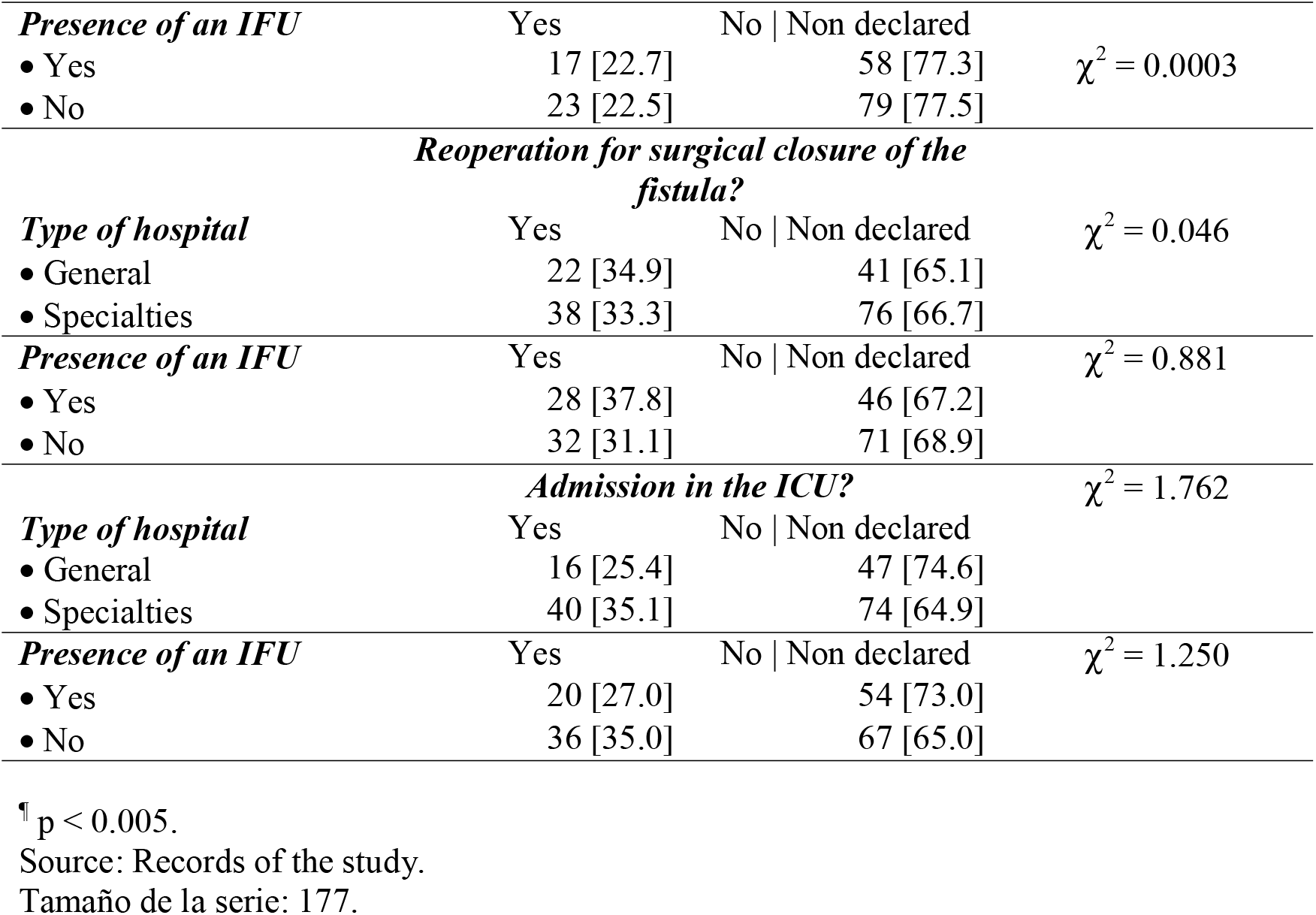
Influence of the operational characteristics of the participating hospital upon the availability of the assessed surgical practices.

Existence of a IFU within the participating hospital did not influence either upon the surgical practices conducted for containing GIF. An exception is to be made with the technique of open abdomen (*IFU present*: 20.3 % vs. *IFU absent*: 34.0 %; Δ = -13.7 %; p < 0.05) and the use of a device for temporary closure of the abdomen (*IFU present*: 22.9 % vs. *IFU absent*: 43.1 %; Δ = -20.2 %; p < 0.05), that were less used in those hospitals incorporating an IFU within their organigram.

### Influence of the compliance with guidelines for treating gastrointestinal fistulas upon surgical practices

The “Fistula Day” found surgical practices were conducted in GIF patients in agreement with an specified guideline in 62.0 % of the instances. The most used guidelines were (in descending order): *ESPEN Guidelines*: 33.6 %; *FELANPE Guidelines*: 30.9 %; *Mexican Guidelines for the treatment of the hostil abdomen*: 10.9 %; and *SOWATS Guidelines*: 3.6 %; respectively. Nineteen-point-one percent of the GIF patients were treated with other guidelines not included in those previously mentioned.

The “Fistula Day” explored the influence of the compliance of medical teams with an specified guideline upon evolution and outcome of GIF. Compliance with a guideline did not influence upon survival of the GIF patient: *Compliance with guideline*: 17.3 % vs. *No compliance with guideline*: 10.4 % (Δ = +6.9 %; p > 0.05). On the other hand, compliance with the guideline determined a higher risk of refistulization in the GIF patient: *Compliance with guideline*: 18.2 % vs. *No compliance with guideline*: 7.5 % (Δ = +10.7 %; χ^2^ = 3.945; p < 0.05); and prolongation of hospital stay: *Compliance with guideline*: 54.5 % vs. *No compliance with guideline*: 35.8 % (Δ = +18.7 %; χ^2^ = 5.85; p < 0.05).

Compliance of medical teams with guidelines did not determine the behavior of the cohort of cases either. Compliance with guideline only meant a higher refistulization rate in each time point of the cohort: Second time point: *Compliance with guidelines*: 8.2 % vs. *No compliance with guidelines*: 1.5 % (Δ = +6.7 %); Third time point: *Compliance with guidelines*: 10.9 % vs. *No compliance with guidelines*: 6.1 % (Δ = +4.8 %; χ^2^ = 6.72; log-rank test; p = 0.05). Although the effect was marginal, it is striking given the observational nature of the present study.

Compliance with guidelines did not influence upon the behavior of the cohort of cases after desaggregating it regarding either the condition of the GIF patient upon discharge or prolongation of hospital stay.

Finally, Table 3 shows the associations between compliance with guidelines for treating GIF and surgical practices conducted for their containment. Conducted surgical practice was independent from compliance of the medical team with the selected management guideline (data not shown).

**Table 3.** Associations between compliance with guidelines for treating gastrointestinal fistulas and surveyed surgical practices.

| Surgical practice | Compliance with guidelines |  | Interpretation |
| --- | --- | --- | --- |
|  | Yes | No \| Non declared |  |
| Size | 110 | 67 |  |
| ***CAT + oral ingestion of contrast*** | | | $\chi^2 = 1.334$ |
| • Yes | 46 [41.8] | 34 [50.7] |  |
| • No | 64 [58.2] | 33 [49.3] |  |
| ***Open Abdomen*** | | | $\chi^2 = 0.178$ |
| • Yes | 36 [32.7] | 24 [35.8] |  |
| • No | 74 [67.3] | 43 [64.2] |  |
| ***Use of a device for temporary closure of the abdominal wall*** | | | $\chi^2 = 0.387$ |
| • Yes | 36 [32.7] | 25 [37.3] |  |
| • No | 74 [67.3] | 42 [62.7] |  |
| ***Use of somatostatin / analogs*** | | | $\chi^2 = 0.0006$ |
| • Yes | 36 [32.7] | 25 [37.3] |  |
| • No | 74 [67.3] | 42 [62.7] |  |
| ***Reoperation for closure of the fistula*** | | | $\chi^2 = 0.025$ |
| • Yes | 44 [40.0] | 26 [38.8] |  |
| • No | 66 [60.0] | 41 [61.2] |  |
| ***Admission to ICU*** | | | $\chi^2 = 0.418$ |
| • Yes | 39 [35.5] | 37 [55.2] |  |
| • No | 71 [64.5] | 50 [44.8] |  |

Source: Records of the study. Tamaño de la serie: 177.

## DISCUSSION

The present report extends and complements the results shown in previous publications on the current state of containment and treatment of GIF in LATAM hospitals.^5-6^ In the present occassion surgical practices conducted in GIF patients during hospitalization were examined. Surgical practices were conducted only in a third of the patients, but they concentrated among those patients with an EAF. Of the examined surgical practices, use of the technique of open abdomen and reoperation for closure of GIF were associated with prolongation of hospital stay and refistulization, but without further influence upon patient’s mortality. Absence of a definitive effect of a given surgical practice upon evolution and outcome of GIF might seem to depend upon (to a large extent) the type of GIF, as it was corroborated by means of techniques of logistic regression.

Conduction of surgical practices upon containment and resolution of GIF does not seem to depend upon the operational characteristics of the participating hospital either. In fact, surveyed surgical practices were less used in those hospitals having an IFU. For the same reason, conduction of surgical practices was also independent from compliance of medical teams with an specified guideline for the managment of GIF.

Compliance of medical teams with an specified GIF guideline did not mean a better treatment and resolution of this condition either. On the contrary: compliance with guidelines was associated with prolongation of hospital stay and a higher rate of refistulization.

The observational nature of the present research impedes the explanation of the causes for the current state of the affairs. Surgical practices as examined during the “Fistula Day” are always mentioned as those ones determining a better management of GIF, and thus, a better containment and resolution of GIF. It is then only counterproductive administration of these surgical practices to produce the opposite effect, and be followed by a higher rate of refistulization and prolongation of hospital stay, these events determining in turn the increase of costs of medical care and a higher use of highly demanding medical technologies such as mechanical ventilation and admission in the ICU.

Type of GIF seems to determine the effect of the surgical practice administered to the patient, as it was corroborated with the use of logistic regression. It also seems surgical practices are administered to the patients regardless the type of GIF, and their likely capability to benefit from them.

Imagenological visualization of the fistula course is essential for elaboration of prognostic judgements about the likely closure of the fistula.^13^ It does not seem medical teams have difficulties in their access to imagenological techniques when the rate of usage of these resources was independent from the level of the hospital within the health system as well as from the presence of a hospital IFU.

Techniques of open abdomen and use of a specified device for temporary closure of the abdominal wall might be indicated on those surgically complex cases in order to contain the abdominal damage, to estabilize the humoral and clinical condition of the GIF patient, and to approach better the management of GIF. However, it does not seem these practices to be of universal use in any setting and with every type of fistula, given the fact their use did not associate with a better outcome. Besides, the implementation of such solutions implies a closer and more intensive follow-up of the GIF patient, and hence, a higher use of hospital resources.

Use of somatostatin (or their analogs) as a pharmaceutical agent promoting closure of GIF was fostered in the past.^14^ The present work revealed somastotatin and their analogs were hardly used in one-fifth of GIF patients. However, use of these agents was higher in EAF: a counterproductive solution, unless their use was justified to reduce the fistula output. Availability of somatostatin was independent from the operational characteristics of the hospital and the existence of a hospital IFU. In the end, use of somatostatin and their analogs did not determine a better outcome of GIF.

It is discussed if reoperation might serve to speed up the closure of GIF. In this work reoperation was associated with a high rate of refistulization and prolongation of hospital stay. For several authors, postponing the decision about the proper time for surgical closure might mean a lower recurrence of GIF. However, adoption of a conservative position on the closure of GIF must be reconciled with hospitalization time of the patient, available hospital resources, and costs of medical-surgical care.^15-16^ In this regard, Christensen *et al*. (2021)^17^ have reported results similar to those ones exposed by the “Fistula Day”. Thus, rate of post-operatory complications was high in the desscribed serie at the expenses of local | systemic events of sepsis and suture dehiscence.^17^ Although dehiscence of intestinal suture was treated with an emergency surgery, this practice resulted in a seven-fold increase in mortality.^17^ However, and in constrast with the results described by Christensen *et al*. (2021),^17^ the authors of the present work found type of surgery performed for GIF resolution only affected length of hospital stay.^5^

The high surgical, microbiological and metabolic risk of GIF patients should lend to the construction of hospital settings, and the endowment with technological resources, required foe their management, such as the ICU. Availability of hospital ICU was independent from the operational characteristics of the hospital and/or the presence of an IFU. However, use of ICU only meant a higher risk of mortality for the GIF patient. Given the observational nature of the present study, a plausible explanation for this finding can not be advanced. It can only be said clinical, surgical and metabolic course of GIF patients has gone beyond a critical point actions seen as “heroic” such as admission in the hospital ICU might not serve to reverse an ominous outcome.

The present work has also shown medical teams for care of GIF patients do not exploit maximally those operational characteristics of the participating hospitals that would serve for a better management of GIF such as the IFU. Among other roles, IFU might serve as an advisor in the use of techniques aimed to contain and resolve GIF, as it has been suggested.^18^ As a matter of fact, the present work revealed use of techniques of open abdomen and of devices for temporary closure of abdominal wall was lower in those hospitals where an IFU was operative, lending evidence to its involvement in GIF management. In this regard, a previously published essay on the influence of the hospital upon containment and resolution of GIF showed existence of an IFU translated to a higher likelihood of spontanoeus closures of GIF (that is: without the need for reoperation).^6^ It is then likely medical teams, assisted by the hospital IFU, might adopt a conservative attitude in the management of GIF, thus avoiding “heroic” measures for achieving closure of GIF.

## Data Availability

All data produced in the present work are contained in the manuscript. Authors encourage potential readers to submit their proposal for data reviewing and sharing

## CONCLUSIONS

Currently, actions foreseen for the containment and resolution of GIF are conducted in less than half of the patients diagnosed with this condition, regardless the type of fistula, without translating to a better outcome. Although most of the medical teams follows a specified GIF management guideline, surveyed surgical practices are conducted independently of the norms prescribed in the guidelines.

### Further extensions

Nutritional practices adopted by medical care teams during containment and resolution of GIF will be examined in an upcoming work. Use of one or other surgical practice in GIF patients is not enough if they are not accompanied by a bundle of nutritional care. Being GIF a hypercatabolic and cachectizing event, nutritional therapy shoud be a comprehensive part of hospital care in order to achieve the sinergies required for control of sepsis, inflammation and resistance to insulin; attenuation of tissue catabolism and promotionof tissue healing and accretion.

## Footnotes

* Available at: http://www.redcap.org.

